# Was access and quality of healthcare affected during COVID-19 pandemic? A qualitative enquiry into healthcare access for non-communicable diseases in Central India

**DOI:** 10.1101/2023.02.23.23286390

**Authors:** Raunaq Singh Nagi, Anirban Chatterjee, Kritika Singhal, Arun M Kokane

**Author notes:** **Corresponding author:** Anirban Chatterjee, Affiliation and postal address: Department of Community and Family Medicine, All India Institute of Medical Sciences, Bhopal, Madhya Pradesh, India, E-mail ID. Postal Address: Department of Community and Family Medicine, All India Institute of Medical Sciences, Bhopal, Madhya Pradesh – 462024, India. e-mail ID.

## Abstract

**Objective:** COVID-19 pandemic has had significant impacts on healthcare systems across the world. However, its impact on healthcare systems in Low- and Middle-Income Countries (LMICs) has been especially devastating, resulting in restricted access to healthcare. The present study was conducted to assess healthcare access for non-communicable diseases (NCDs) in Central India.

**Design:** Inductive and deductive thematic analysis of in-depth semi-structured interviews.

**Setting:** Study was conducted in communities of two urban and rural districts of central India.

**Participants:** Interviewed participants included PLNCDs, their caregivers, community dwellers, CHWs such as, Accredited Social Health Activists (ASHAs) and Anganwadi Workers (AWWs), Medical Officers, and Community Leaders. Recruitment of the participants was done via purposive and convenience sampling.

**Result:** A total of fifty Key Informant Interviews were (KIIs) conducted. All participants reported facing considerable difficulties while trying to access care from both public as well as private healthcare facilities. Absence of staff, equipment and medicines, restricted commute, misconceptions regarding the spread of COVID-19, and the stigma attached to COVID-19 infection acted as major barriers to accessing care, while door-to-door visits by community health workers, community support, and presence of privately owned healthcare facilities in the vicinity acted as facilitators.

**Conclusion:** In our study, we found that continued functioning of primary healthcare centres, ensuring uninterrupted supply of medicine and effective dissemination of information regarding COVID-19 could have acted to ease access to healthcare. Going ahead, capacity building to offset the impact of future emergencies and pandemics should be a crucial consideration while developing resilient healthcare systems.

**Strengths and limitations of this study:**

- Our study is the first study to explore the barriers faced by PLNCDs of low socio-economic status during the pandemic.
- We explored the perspectives of both patients and healthcare workers before triangulating the data findings.
- The study was conducted in the PLNCDs of lower socio-economic group and hence the perspectives and experiences of other socio-economic groups are yet to be explored.

## Introduction

Non-communicable diseases (NCDs) kill 41 million people globally each year – equivalent to 71% of total deaths globally. Seventy-seven percent of these deaths occur in Low- and Middle-Income Countries (LMICs) (1). In India, 62% of all deaths is accounted for by NCDs, with NCDs being four of the five leading individual causes of Disability Adjusted Life-Years (DALYs) in 2016 (2). However, morbidity and premature mortality arising as a result of NCDs can be controlled by implementing preventive strategies focusing on lifestyle modification and awareness management, along with ensuring early detection via active screening and continued care of NCDs (3).

The ongoing COVID-19 pandemic has severely impacted healthcare systems across the globe. Studies across multiple health systems have consistently documented a dip in the utilization of healthcare services coinciding with the COVID-19 pandemic outbreak and consequent mobility restrictions (4,5). A systematic review on the impact of COVID-19 pandemic on healthcare services found that utilization of healthcare services during the pandemic reduced by as much as one third across healthcare systems (6).

Health systems in LMICs are usually very fragile and non-resilient. Access to healthcare outlets continues to be an issue with most health systems across LMICs (7). Public healthcare spending across LMICs is comparatively low, resulting in high out-of-pocket expenditure (8). In addition, the ratio of healthcare staff and hospital beds to patients is usually very low (9). As a result, the COVID-19 pandemic resulted in large scale disruptions within health systems in LMICs.

NCD management is highly dependent on regular assessment and medication of people living with NCDs (PLNCDs). However, the COVID-19 pandemic interrupted service delivery to these people. A rapid survey by WHO assessing NCD access in the wake of COVID-19 pandemic and associated restrictions found that access to outpatient NCD services was interrupted in 59% countries (10). Studies from India (11) and other LMICs have also found decreased utilization of NCD services with the advent of COVID-19 (12,13). However, there is limited information on the impact of COVID-19 on NCD healthcare provision and access from India. In the current study therefore, we have qualitatively explored the barriers and facilitators experienced by patients with NCDs while trying to access healthcare services from Bhopal and Raisen districts of Madhya Pradesh – a state in central India.

## Methods

Semi-qualitative interviews were conducted amongst 50 stakeholders from various sections of the society and the healthcare delivery system. Data was collected from February, 2021 to April, 2021 from both rural and urban Madhya Pradesh. Data from rural Madhya Pradesh was collected from Community Health Centre (CHC), Goharganj block, Raisen district, while that from urban Madhya Pradesh was collected from Urban Primary Health Centre (UPHC), Sai Baba Nagar urban slum and the ambulatory out-patient department (OPD) of All India Institute of Medical Sciences (AIIMS) Hospital, Bhopal. In addition, we also contacted FLWs in our catchment areas to enquire about PLNCDs in the community who are not availing services at the abovementioned healthcare centres. Interviewed participants included PLNCDs, their caregivers, community dwellers, CHWs such as, Accredited Social Health Activists (ASHAs) and Anganwadi Workers (AWWs), Medical Officers, and Community Leaders. The healthcare centres had pre-existing line lists of PLNCDs. Recruitment of the participants was done via purposive and convenience sampling in order to ensure adequate representation of all stakeholders. Interviews were conducted at mutually convenient venues – OPDs, village community buildings, or at the residence of the participant – as agreed upon by the interviewer and the participant. PLNCDs were often accompanied by caregivers, though privacy was ensured for the duration of the interview. Details of the study population are presented in Table 1.

**Table 1:** Characteristics of study population

|  | PLNCDs | Caregivers | Community Dweller | ASHA | Medical Officer | ANM | Anganwadi Worker | Community leader | Total |
| --- | --- | --- | --- | --- | --- | --- | --- | --- | --- |
| Rural | 4 | 5 | 1 | 5 | 1 | 1 | 0 | 1 | 18 |
| Urban | 27 | 0 | 0 | 3 | 0 | 0 | 2 | 0 | 32 |

Prior to initiation of interviews, informed consent was obtained from all the participants and they were also provided with information related to the study and their role in it via a leaflet in Hindi and English. All the interviews were conducted with the help of pre-tested/piloted interview guides written in the vernacular language (Hindi). The interview guide included questions eliciting information about the participants’ socio-demography, the barriers they had to negotiate with while accessing NCD healthcare during COVID-19 lockdown, the facilitators which eased their access to healthcare services, and measures which can be implemented to ease access to healthcare in case similar restrictions are placed in the future. All the interviews were conducted face-to-face while maintaining all COVID-19 relevant protocols.

The interviews were conducted by RSN (male) and KN (female), and audio recorded using a handheld recorder. They were initially transcribed verbatim into Hindi, and were then translated into English. Neither the verbatim transcripts nor translations were not returned to the participants for verification. Interviews were analysed by three researchers (RSN, AC and KS) independently using the constant comparative method to derive at codes, sub-themes and themes from within the textual information. Although a few themes had been determined *a priori*, interpretative phenomenological analysis using thematic approach was rigorously applied to derive novel themes also. After analysing the first few transcripts separately, they developed code books which were compared and condensed into a final code manual. This was used for analysing rest of the transcripts. The researchers analysed the interviews independently, and once coding was complete, the team (all authors) met to discuss and refine emergent themes. NVivo 12 plus was used by the researchers in order to develop a matrix of the perceived barriers, facilitators, and measures which can be taken to ease access to healthcare services. Interviews were continued till data saturation was obtained – as evidenced by increasing overlap in the information provided by participants.

Prior to data collection, institutional ethics clearance was obtained from the Institutional Human Ethics Committee (IHEC-LOP/2021/IM0298).

## Results

Fifty stakeholders (Table 1) were interviewed from February to April, 2021. The median time of interviews was twenty minutes (fifteen – twenty-seven minutes). All the stakeholders who were approached for soliciting participation in the study consented for the same.

Table 2 presents the themes and subthemes of our findings. We found two themes, with seven sub-themes in total. Three sub-themes pertained to all three domains, while two sub-themes were related to two domains (one for barriers and facilitators, and one for barriers and desired actions). Two sub-themes pertained to one domain only. The sub-themes have been diagrammatically represented in Fig 1.

**Table 2:**
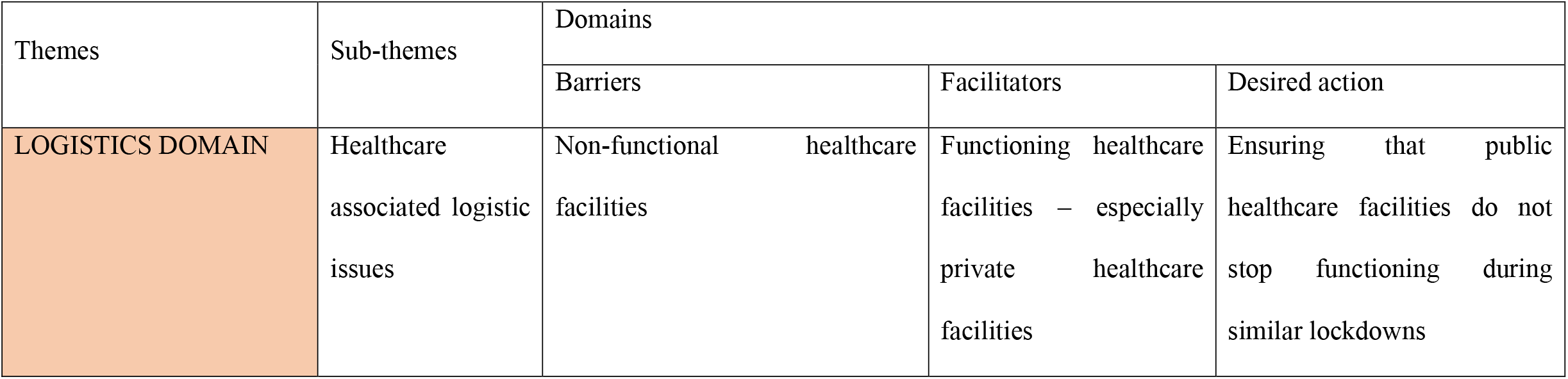
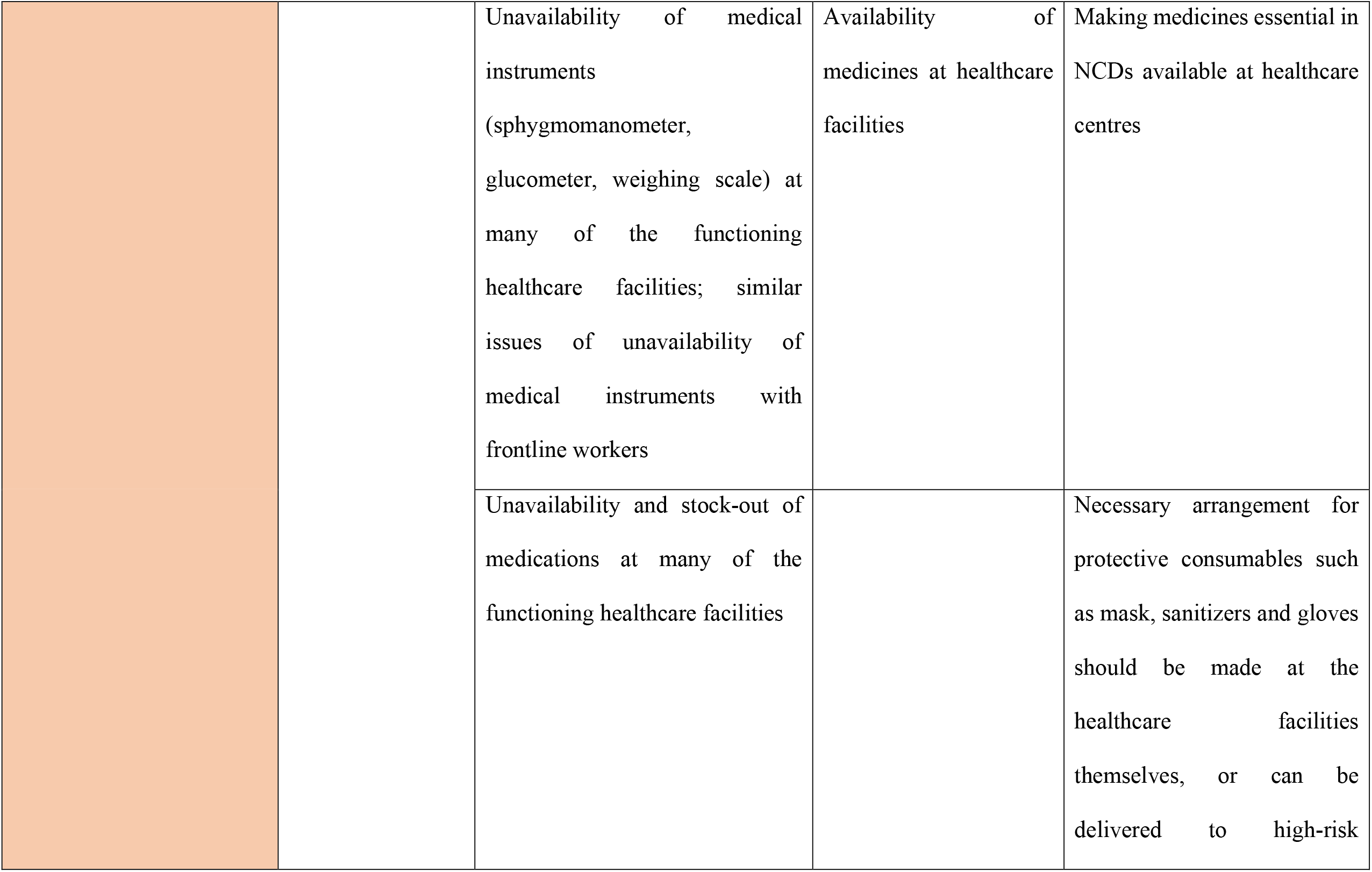
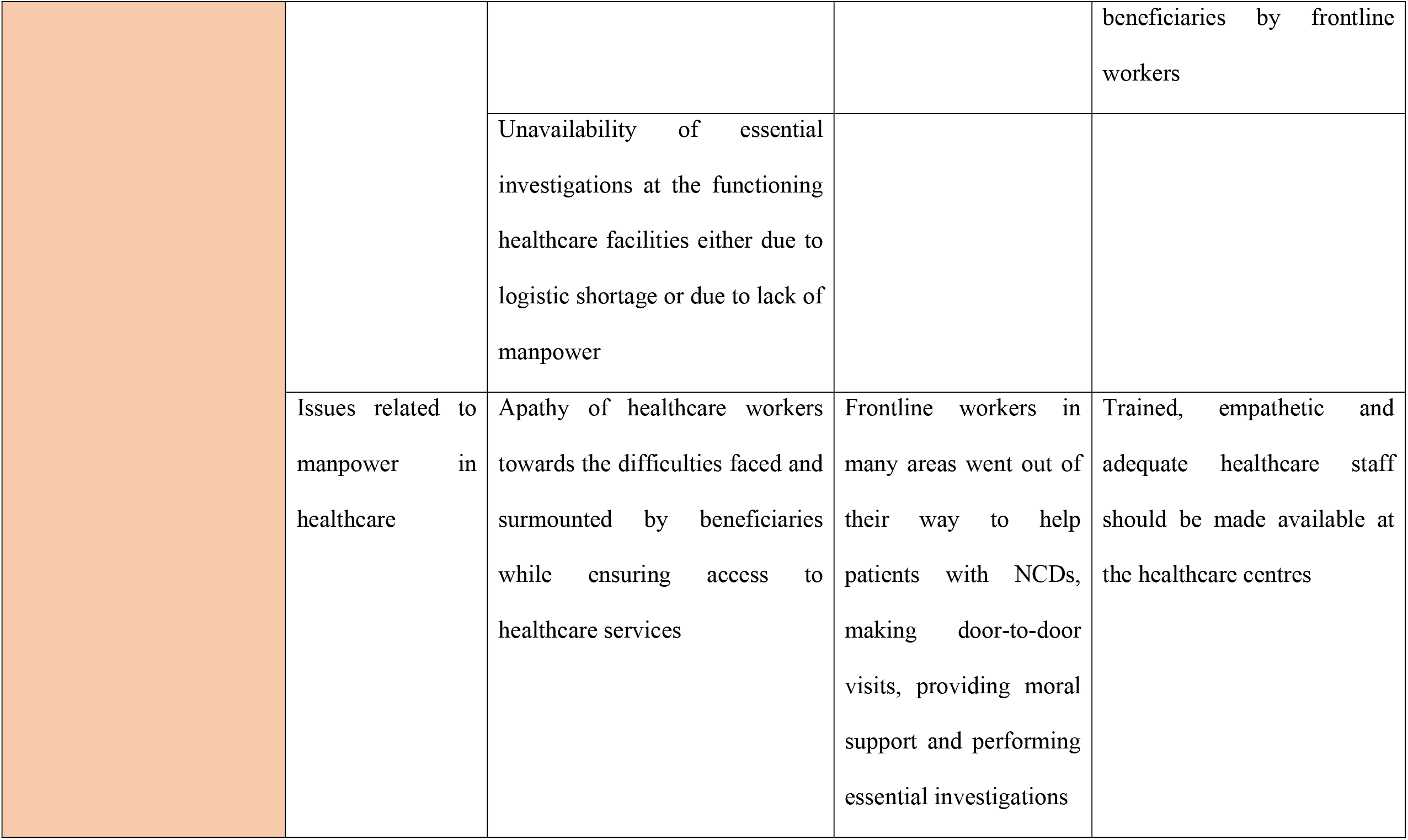
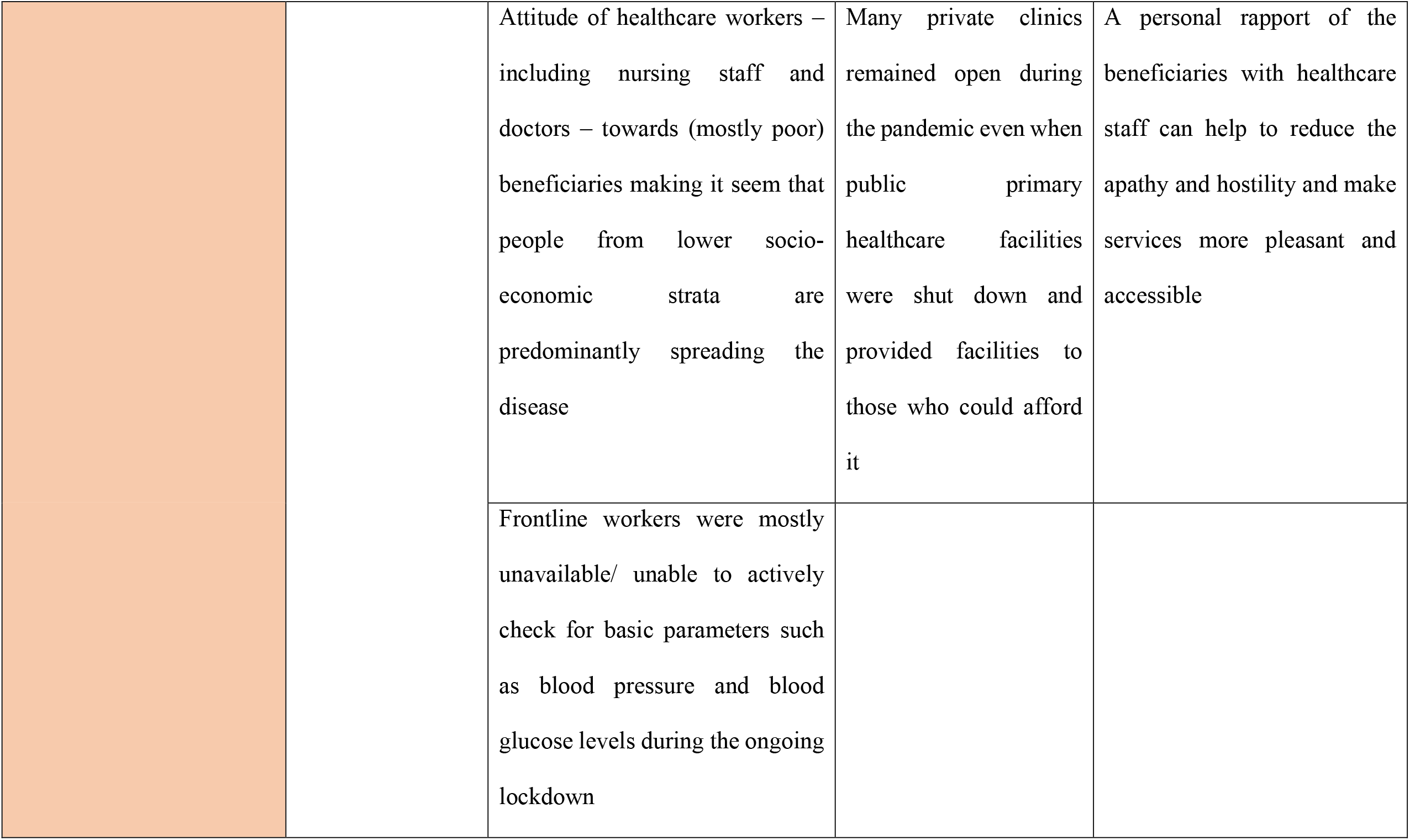
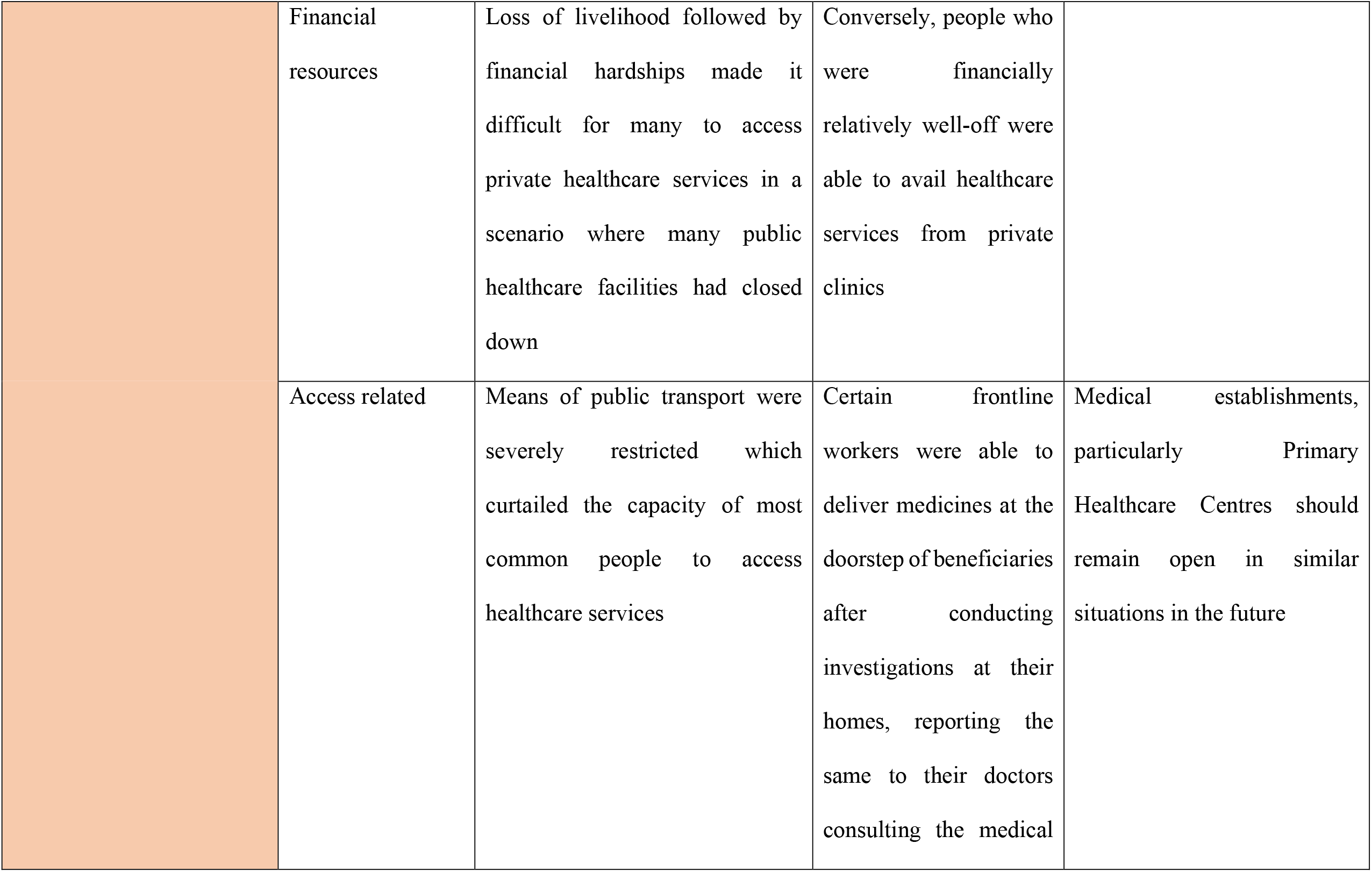
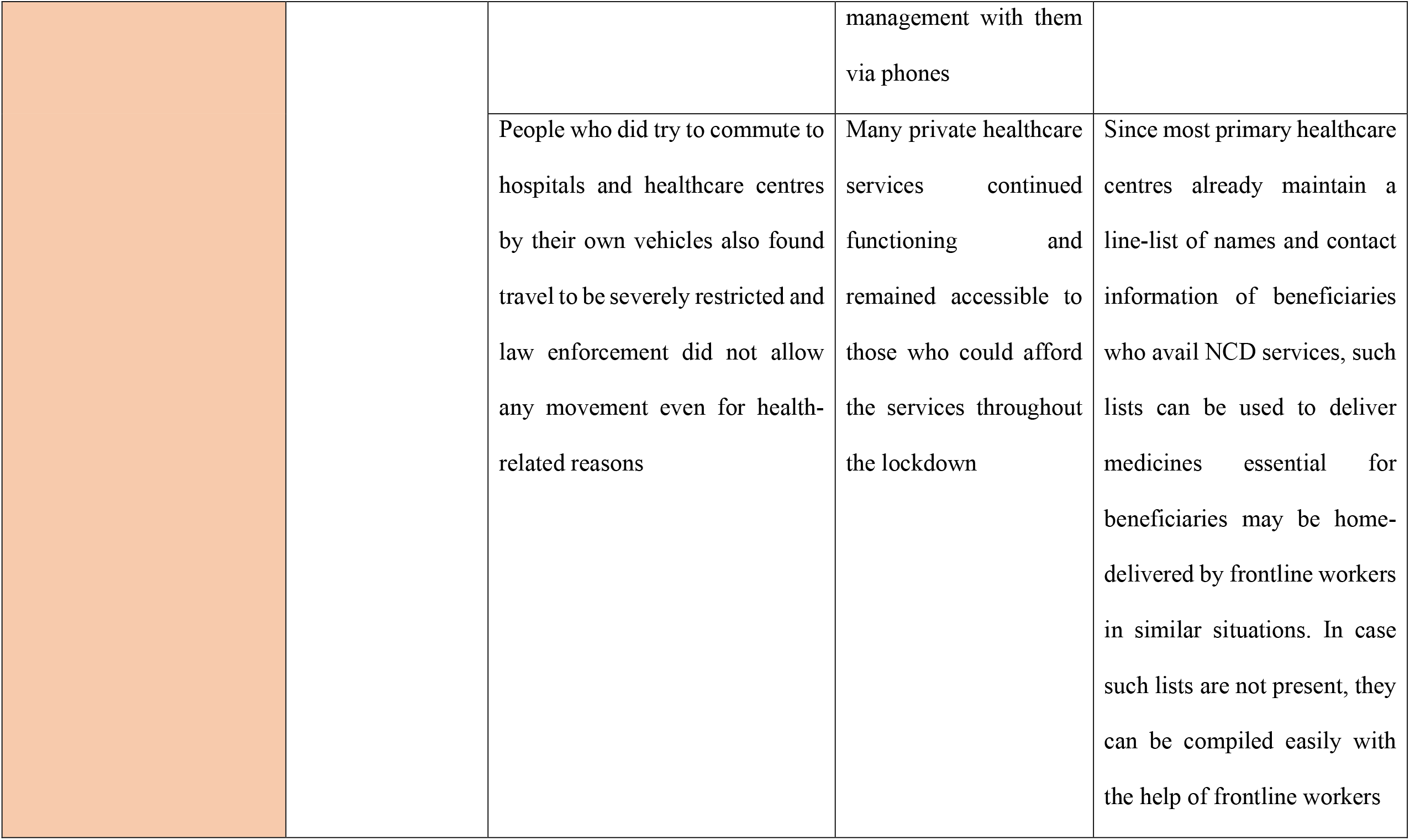
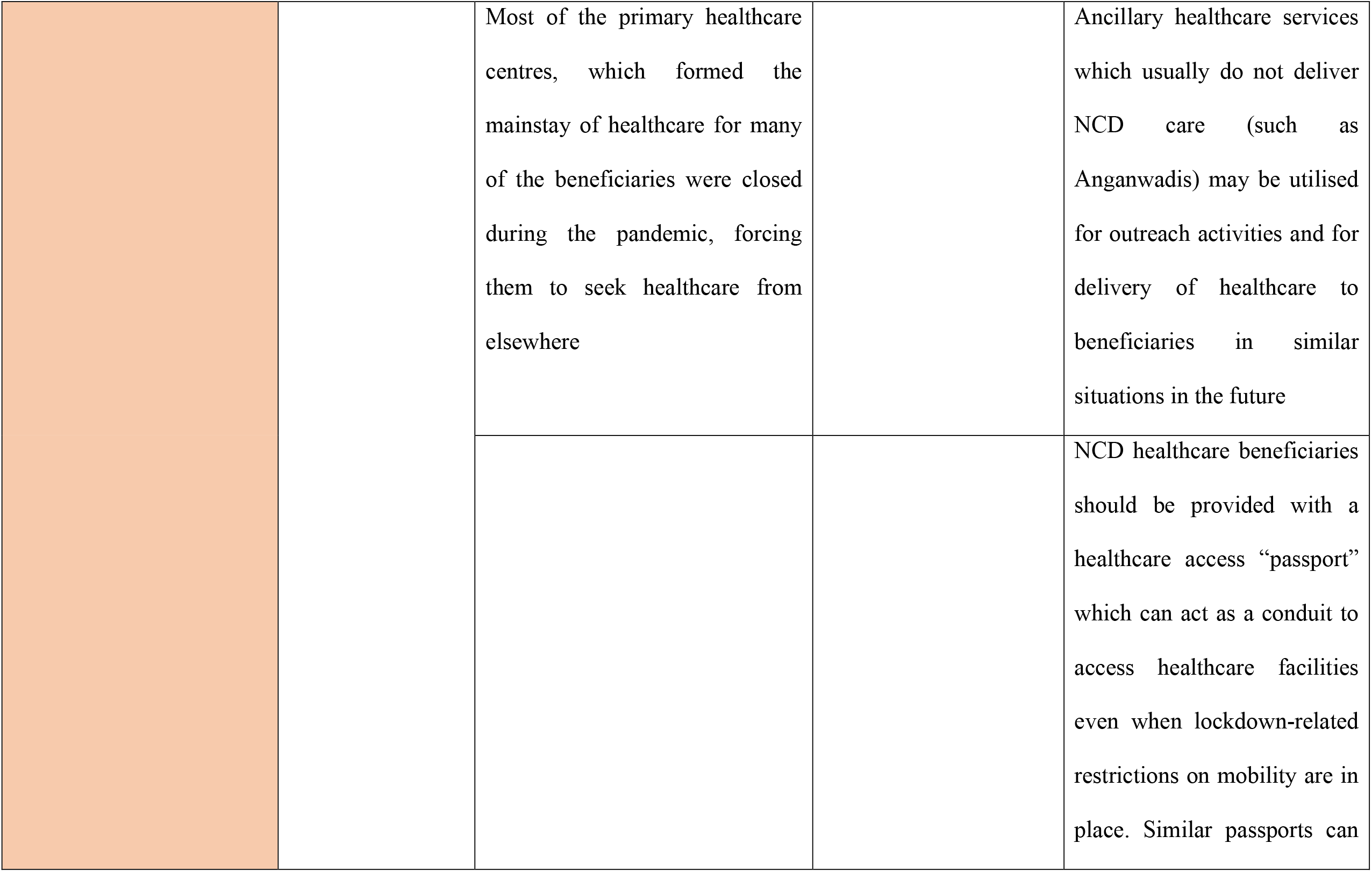
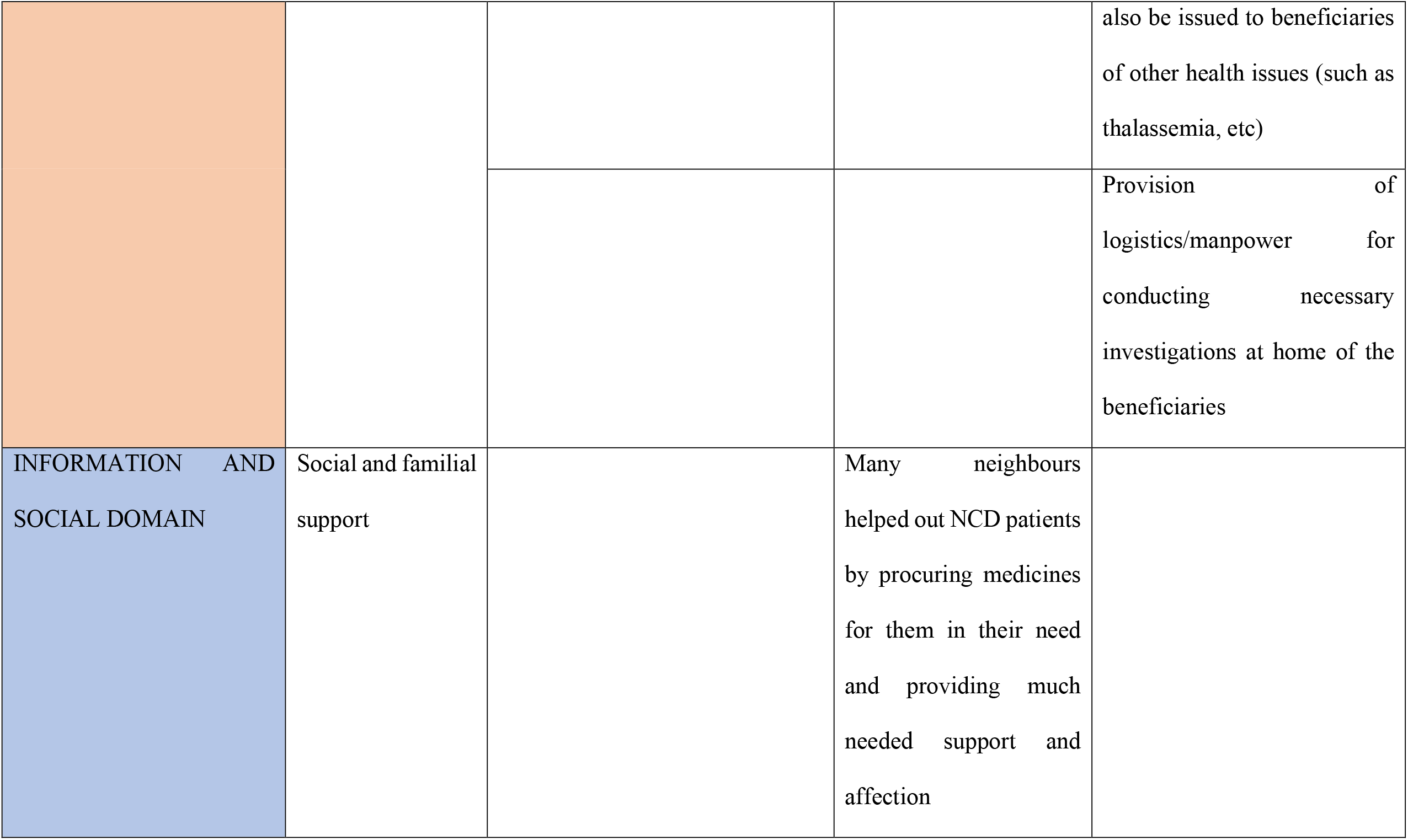
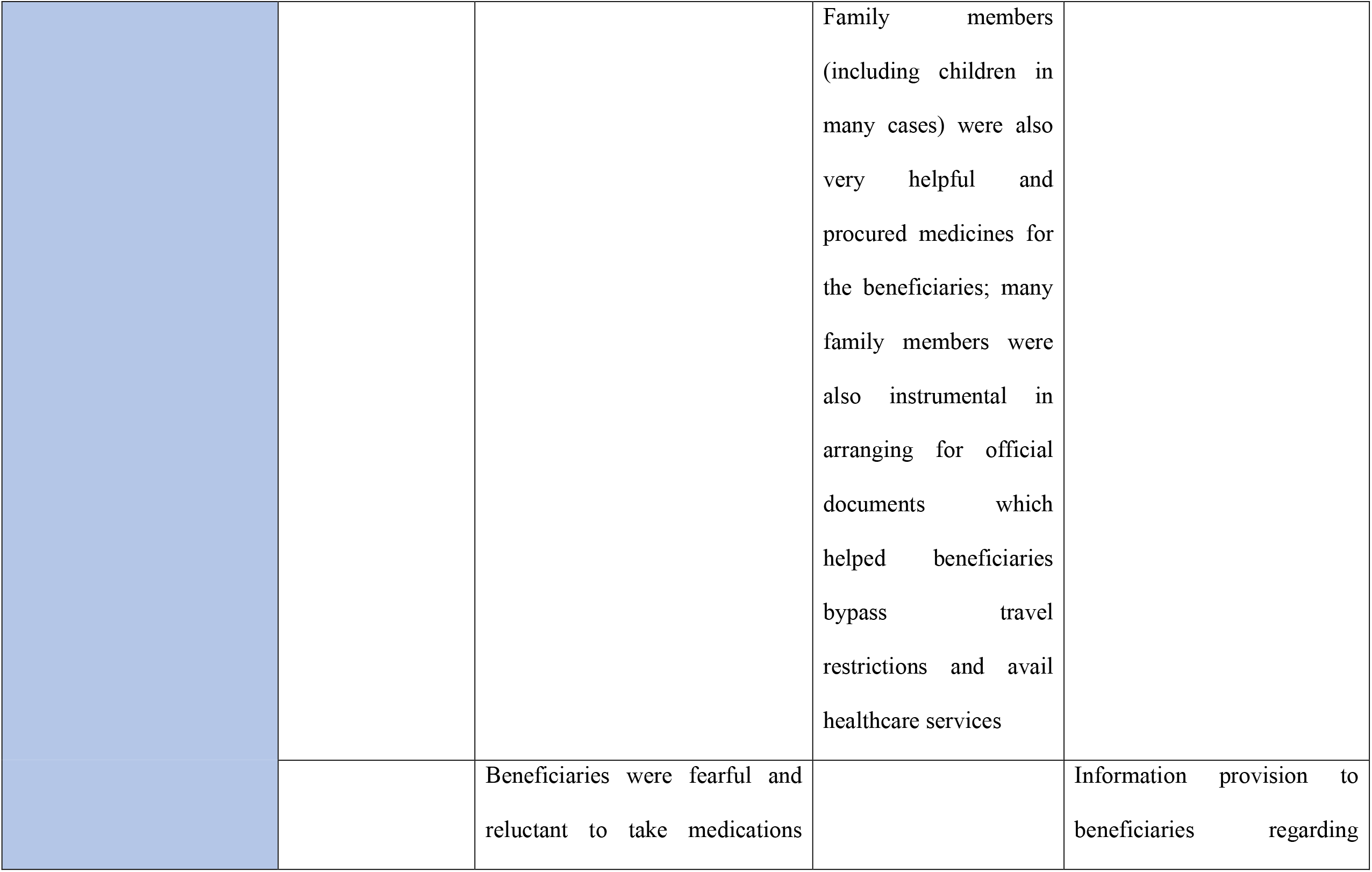
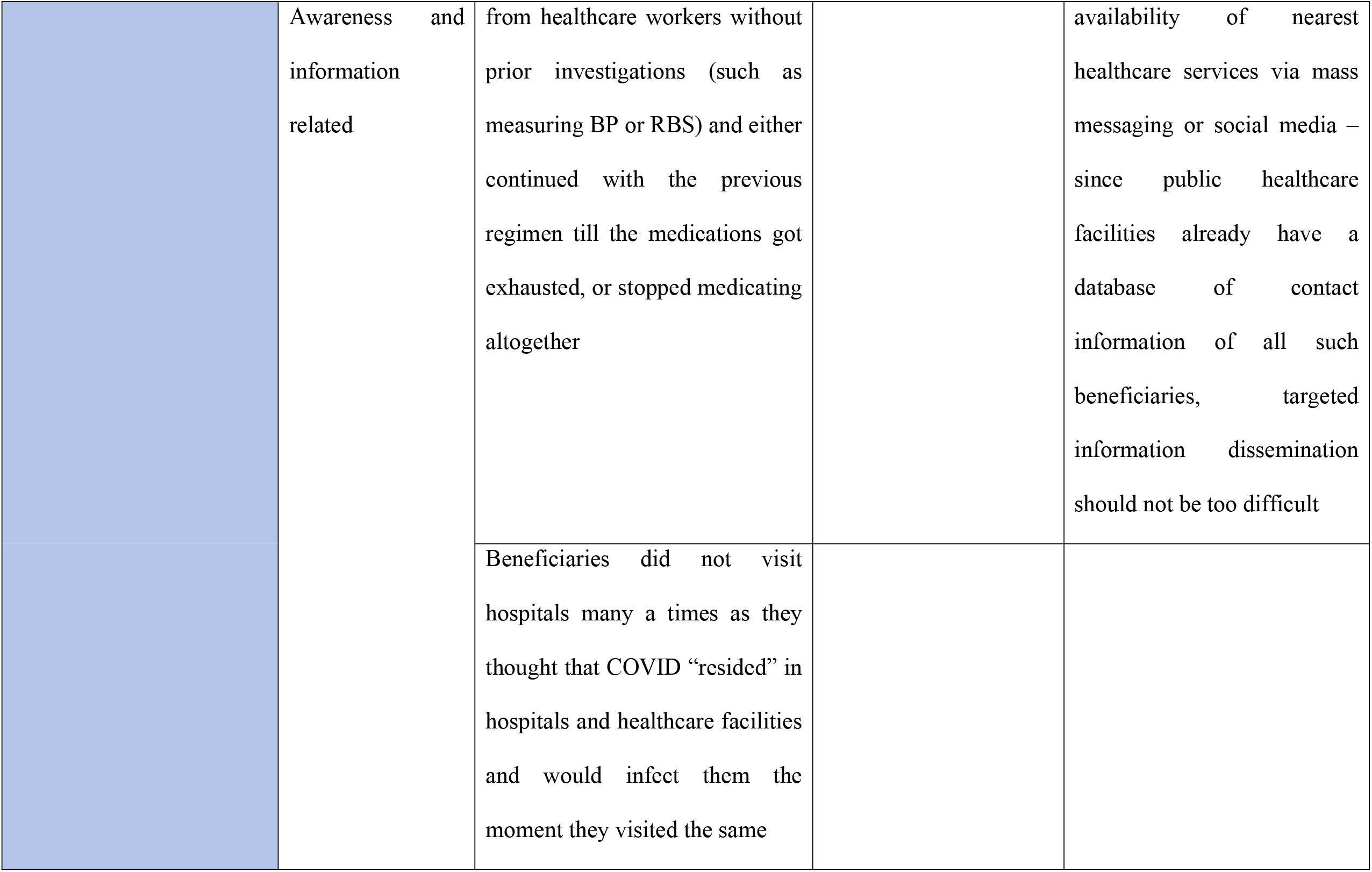
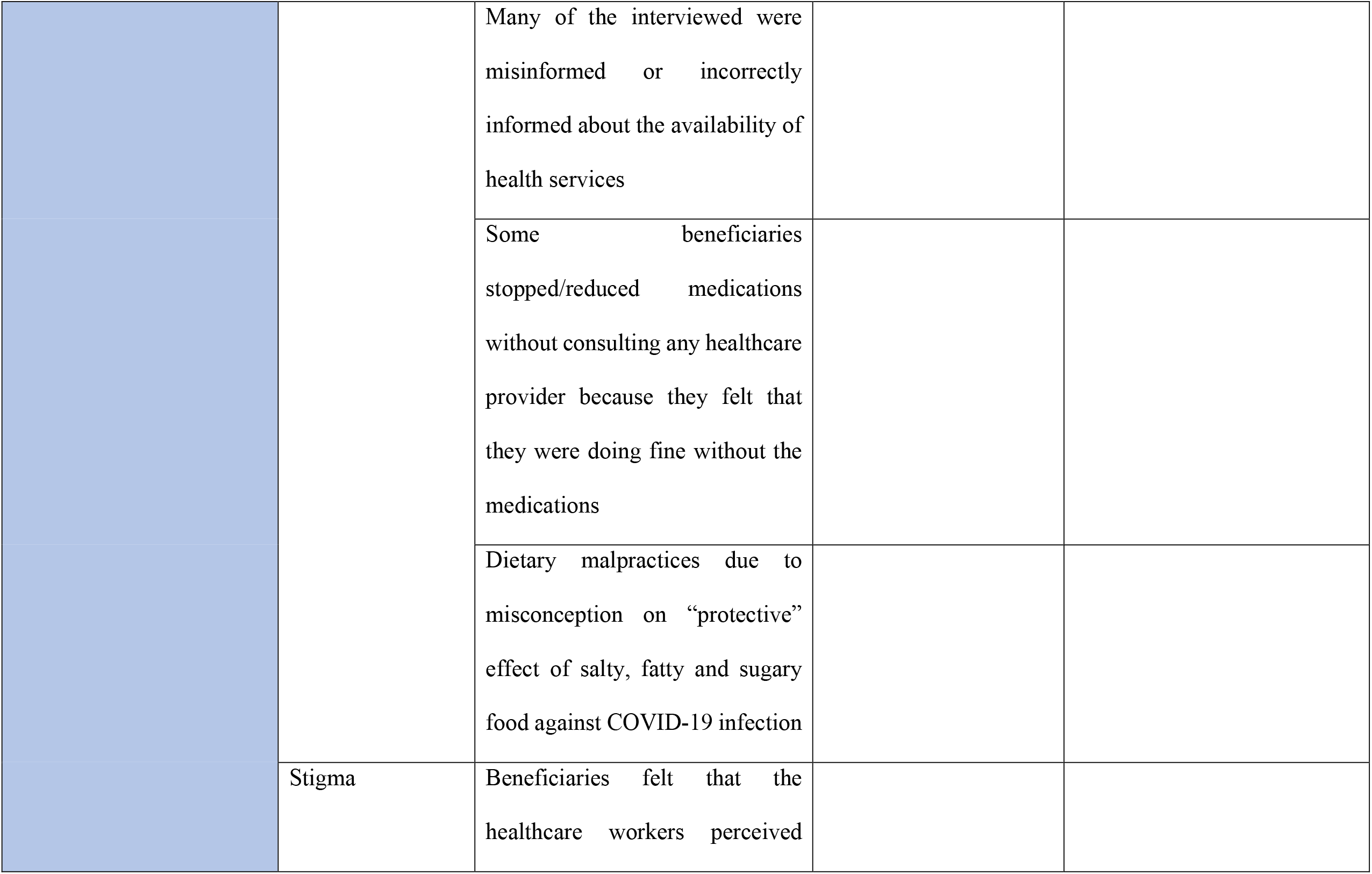
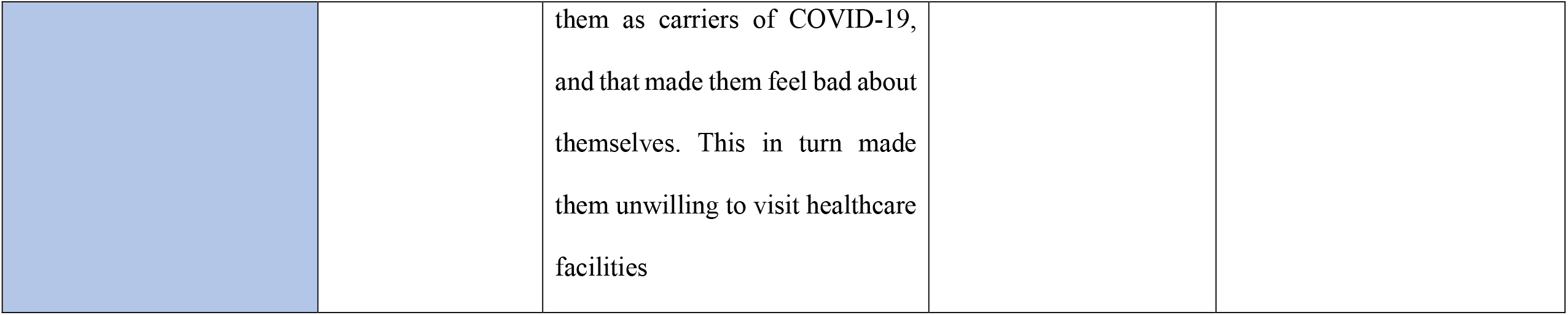
Identified barriers and facilitators associated with access to healthcare by NCD patients during the COVID-19 associated lockdown and desired actions for improving access to healthcare services in similar situations in the future.

**Fig 1:**
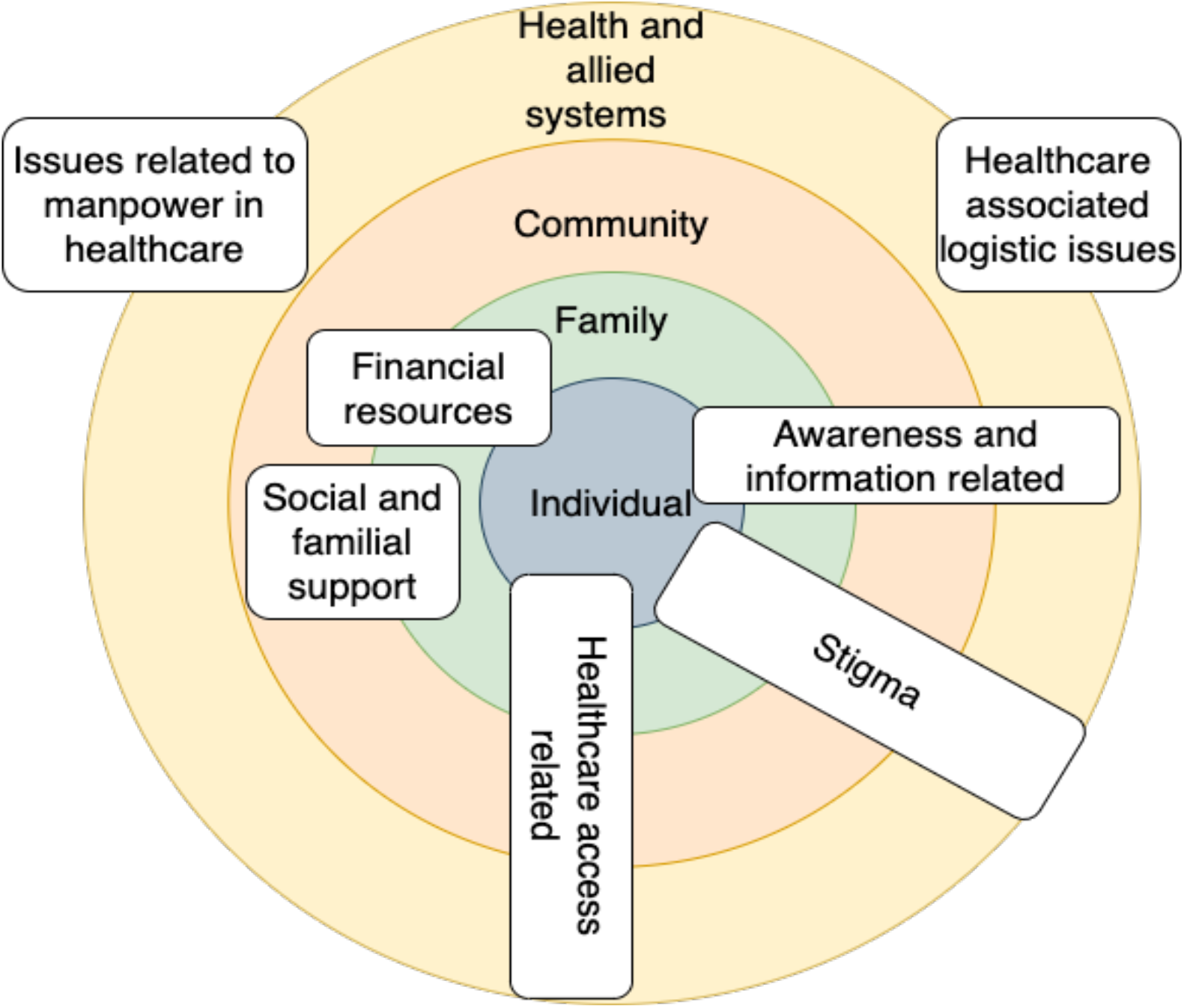
Diagrammatic representation of determinants of access to healthcare systems. *Some determinants such as stigma and healthcare access related span from the individual to systemic domains, while some determinants such as social and familial support and issues related to manpower in healthcare only span a few of the domains*.

### Themes

#### Healthcare system related

Several factors pertaining to healthcare system were identified to function as barriers and facilitators in the NCD care of the beneficiaries.

##### 1. Healthcare staff and doctors

Healthcare facilities were reported to be mostly non-functional during the pandemic, which hindered routine services pertaining to NCD care. In the rare case that a healthcare facility was open, it did not have any medical officers (duty doctors) as they had been absorbed into tertiary care of COVID. This severely impeded access to healthcare.

On the other hand, some beneficiaries were able to access functional healthcare facilities and outreach activities. However, most such facilities were privately owned and run. Outreach activities in the form of door-to-door visits for communicating COVID appropriate behaviour and guidelines facilitated a better understanding of the situation and also eased access to healthcare services by the beneficiaries, as such visits were usually conducted by frontline workers, who were able to ensure continued medication.

##### 2. Medicines and investigations

Several beneficiaries reported unavailability of medical equipment such as sphygmomanometer and glucometers for regular investigations, as they had purposed for providing for tertiary healthcare needs in the face of increased hospitalizations.

Access to routine medication was also hampered. Many beneficiaries reported that they had to resort to alternative means for procuring their routine medication – which included purchasing from often distant pharmacies or taking unidentified “substitute” drugs. Few discontinued their medications as they were unable to access medicines, while some reported that they reduced the dosage to half to make the medications last longer.

A few beneficiaries were able to access medications and medical instruments comparatively easily, but they mostly were able to do so from privately run clinics, laboratories and pharmacies. Primary healthcare services were mostly unavailable.

#### Human resources in healthcare

##### 1. Role of frontline/community health workers

Community Health Workers (CHWs) were pressed into providing continued primary healthcare services in many areas, and were able to meet community demands to a large extent. They conducted vital investigations such as checking blood pressure, consulting doctors for patients who were unable to commute and obtain medicines (such as anti-hypertensive and anti-diabetic medications) and delivering them to the beneficiaries. Presence and cooperation of the CHWs facilitated the access to healthcare by PLNCDs. However, all CHWs were not proactive in providing these services. At least some CHWs contracted COVID-19 themselves and were therefore quarantined. Sphygmomanometers and capillary glucometers which are usually available with CHWs were repurposed to meet increased demands in tertiary healthcare centres. In addition, many CHWs also got posted in tertiary healthcare centres, removing them from community settings.

##### 2. Attitude of healthcare workers and perception of beneficiaries

The interviewed PLNCDs said that they perceived that doctors were discriminating against them. They reported that doctors and healthcare staff were disinterested in their problems and illnesses. Some PLNCDs also reported that they were treated rudely and reprimanded for no apparent reason. It was alleged that doctors and healthcare staff maintained strict physical distance and did not even conduct any physical inspection. Some PLNCDs even reported that their OPD tickets were not being touched, being pushed or thrown away.

Many beneficiaries believed that such behaviour was due to the fear of contracting COVID-19. However, many PLNCDs, especially from the lower socioeconomic strata believed that they were misbehaved with as they were perceived to be more likely to be carrying infection. They in turn felt ‘dirty’ and unsanitary due to the behaviour of the healthcare staff and doctors.

#### Financial resources

Respondents from the lower socio-economic strata reported that the COVID-19 pandemic and the resulting lockdown had led to loss of livelihood, severely limiting their financial means. This, coupled with the unavailability of routine public healthcare services, rendered many relatively poorer PLNCDs unable to access any healthcare at all. Some PLNCDs who were relatively well-off were able to seek and access care from private providers; however, since many private healthcare providers had also ceased their operations, it was an exception, and not the rule.

#### Access related

##### 1. Transportation issues

Many PLNCDs who were dependant on public transportation to commute to healthcare facilities were unable to access care during lockdown as public transportation was not available. Even when beneficiaries were using their private vehicles, reaching healthcare facilities proved to be difficult due to lockdown associated restrictions. In certain cases, law enforcement authorities did not permit movement despite being made aware of medical issues.

##### 2. Access to medicines and services

Many PLNCDs relied on local dispensaries and primary healthcare centres for securing medications. However, many such primary healthcare centres and dispensaries had halted their services during the pandemic. On the other hand, in isolated instances CHWs were able to deliver medications and other healthcare services door-to-door, thereby greatly aiding the beneficiaries. Some pharmacies and private clinics also continued functioning allowing access to medical services to a section of the beneficiaries.

#### Social and familial support

Many respondents informed that social and familial support played a major role in helping them tide over the lockdown and related problems. Not only were friends and neighbours uniformly supportive and understanding, in many cases they put in extra efforts to procure medicines for the interviewed beneficiaries. Similarly, family members were also very supportive and did their best to ensure that they could continue their medical treatment, by trying to procure medications, or by trying to procure documents which would enable beneficiaries to bypass lockdown-related travel restrictions and access healthcare.

#### Awareness and information related

##### 1. Healthcare related misinformation

Many of the respondents reported of multiple misinformation related to the COVID-19 pandemic. Many beneficiaries were reluctant to go to hospitals/healthcare centres, as they thought that the COVID-19 infection “resides” there, and they may get infected. Another common misinformation was regarding the functioning of healthcare facilities. Several of the beneficiaries reported being misinformed about the unavailability of medical staff or closure of healthcare centres. As a result, they avoided necessary medical travel, even when the centres were functional and facilities were functioning.

##### 2. Dietary misconceptions

Some beneficiaries started consuming fat-, salt- and sugar-rich foods under the belief that such “rich” food will strengthen them and protect them from the disease. According to interviewed healthcare workers, this behaviour, coupled with non-consumption of regular medication may have led to further exacerbation of their illness.

Coding tree for the themes is represented in Fig 2. And verbatims are provided in the Supplementary file.

**Fig 2:**
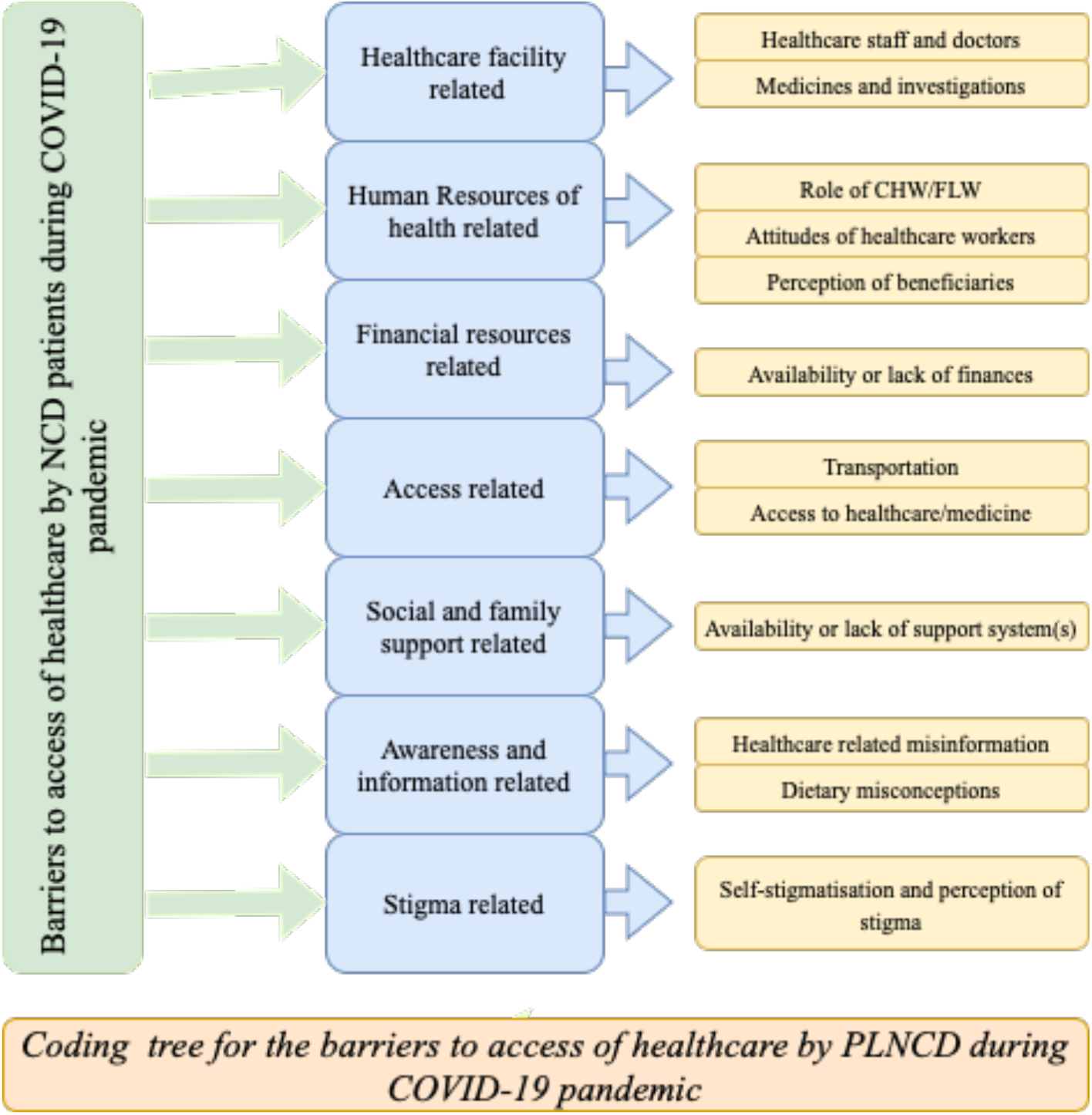
Coding tree for the barriers of access to healthcare services by PLNCD during COVID-19 pandemic. *Representation of the themes and sub-themes of barriers to healthcare access*.

#### Stigma

Many beneficiaries reported that they were made to feel “dirty” by healthcare staff and doctors via their interactions. This made them very resentful, and they felt that they were treated differently or discriminated against. Consequently, such beneficiaries refrained from seeking healthcare – either due to hurt self-esteem, or due to self-stigma and resultant reluctance to interact with others due to fear of transmitting the disease.

### Desired points of action

During the interviews, the participants were requested to list actionable points that could have been implemented during the lockdown, and can be considered for similar future scenarios:

1. Medical establishments – particularly the primary health centres (PHCs) and health and wellness centres (HWCs) – should never be closed during similar pandemics. Similarly, CHWs should also be trained and provided logistic support to ensure uninterrupted provision of medical services and medications. CHWs mentioned that they usually have a list of PLNCDs in their community, which can be employed during similar situations to deliver targeted services, such as investigations and medications. In addition, it was also suggested that a list of nearby operational healthcare centres can be made available with CHWs, so that community members can be directed to the same when needed.
2. Beneficiaries and healthcare workers alike suggested that other facilities, such as *Anganwadi centres* can be used for outreach activities – such as regular health-check-ups for blood pressure and blood sugar or targeted delivery of medicines. *Anganwadi* workers reported that they had been visiting the communities on a regular basis for disseminating messages about COVID associated precautions and COVID appropriate behaviours, in addition to maternal and child care services. They suggested that such visits can also be used for distributing medications.
3. Given that many frontline workers and doctors from PHCs were deputed at tertiary healthcare centres to circumvent the chronic and severe shortage of healthcare staff, one of the most common desired points of action was hiring extra healthcare staff, at least for the duration of the pandemic. Similarly, it was also emphasized that availability of medicines and investigations, particularly in the government set-up should be regularized and uninhibited.
4. CHWs informed that many a times beneficiaries were turned away from healthcare facilities on account of them not having face masks. Given that many PLNCDs are unable to afford face masks, it was suggested that healthcare centres – particularly PHCs and HWCs, should provide masks and sanitizers free of cost or while entering the facilities.
5. Shutting down public transport completely cut off a section of PLNCDs from accessing healthcare. It was suggested that in the future, documents such as NCD drug diary may be used as “passes” allowing them to access public transport services which can be run at limited capacity. Similar “passports” can also be used by beneficiaries while travelling by private vehicles.
6. Finally, both PLNCDs and healthcare workers opined that healthcare staff should be especially sensitised to the needs of vulnerable populations during adversities such as the COVID-19 pandemic, so as to make services and service providers more empathetic and approachable.

## Discussion

Utilization of healthcare services was adversely affected during the COVID-19 pandemic. In one study, it was found that access to healthcare services had decreased considerably during the COVID-19 pandemic – by as much as 33% during the first and second waves of the pandemic (6). This coincided with an increase in all-cause mortality across communities (14). However, people living with NCDs (PLNCDs) suffered from a disproportionately high mortality rate once infected by SARS-CoV-2 (15). Lack of access to healthcare services acted as a double-whammy for PLNCDs – rendering an already vulnerable people even more vulnerable to disproportionately high risk of preventable morbidity and mortality.

In our study, we found that PLNCDs across socio-economic strata faced significant challenges when they tried to physically access healthcare, which is similar to findings in other studies from India (11) and elsewhere (16). Uniquely however, we also found that many a times access was restricted due to lack of financial resources, which rendered securing private consultations and procuring medications out of reach (17).

Availability of healthcare staff was also a major factor in determining access to healthcare services. Ongoing COVID-19 related exigencies resulted in shunting of workforce from primary healthcare to tertiary healthcare (18), and COVID-19 related protocols necessitated reducing in-person consultations and physical examination of beneficiaries. As a result, primary healthcare systems were strapped of human resources. Although efforts were made to integrate telemedicine into primary healthcare delivery systems (19) they were unable to replace conventional primary healthcare in both scale as well as acceptability (20). However, in many healthcare settings frontline workers were pressed into action for ensuring access to routine primary healthcare services with much success (21,22), which was a finding in our study also.

Multiple information-related barriers were also reported by the participants. Pandemic-related misinformation has been extensively circulating on social media (23), and has adversely affected the public health response to COVID-19 pandemic (24,25). However, the success of multipronged intervention packages to combat such infodemics (26) provides evidence for integration of similar strategies in the public health response system for future scenarios.

Self-stigma and social stigma also affected healthcare access. Stigma is a known barrier to healthcare-seeking and social support in multiple social settings (27), during multiple pandemics – both past and present (28,29). Generating and disseminating information focussed around reducing stigma, and inculcating a sense of community in societies and people has been shown to be effective in reducing stigma associated with many health conditions (30).

Our study is one of the few from India which explore the barriers and facilitators associated with accessing healthcare for PLNCDs. Our respondents include care-seekers as well as care-givers from both rural and urban communities, therefore affording a holistic lens on the various determinants of healthcare access. Finally, it also summarizes the expectations and requirements of the care-seekers and care-givers from healthcare systems in similar scenarios for ensuring uninterrupted access to healthcare services.

However, since our study is qualitative, limitations inherent to qualitative studies also apply to our study. In addition, we were mostly able to access information from beneficiaries of the lower socio-economic strata only, thereby mostly excluding the experiences of PLNCDs from comparatively well-off families.

## Conclusion

In our study, we found that people with NCDs encountered a number of barriers while accessing healthcare services. These barriers resulted from underutilization and repurposing of existing healthcare cadre, supply chain disruption and logistic limitations, and confusion and stigma resulting from miscommunication. Going ahead, building resilience in healthcare systems by better utilising existing healthcare cadre and integrating a comprehensive communicative strategy as a core component of public health strategy is imperative to ensure adequate response to similar public health emergencies in the future.

## Supporting information

Supplemental file 1- Verbatims

Supplemental file 2- COREQ checklist

## Data Availability

Data is available from the corresponding author up on making valid request.

## Data availability statement

Data is available from the corresponding author up on request.

## Author controbution

RSN, AC and KS developed topic guides. KS and RSN collected and analysed the data. AC analysed the data and wrote the manuscript. AMK supervised data collection, analysis and report writing. All the authors reviewed and agreed to the final version of manuscript.

## Acknowledgements

The authors would thank all the stakeholders who agreed to participate in the interview which include patients of various NCDs and their caregivers, CHWs-ASHAs, ANMs and AWWs, medical officers and others.

## References

1. World Health Organization. Non-communicable diseases [Internet]. 2022 [cited 2022 Mar 12]. Available from: https://www.who.int/news-room/fact-sheets/detail/noncommunicable-diseases

2. India State-Level Disease Burden Initiative CVD Collaborators, Dandona L, Dandona R, Kumar GA, Shukla DK, Paul VK, et al. Nations within a nation : variations in epidemiological transition across the states of India, 1990 – 2016 in the Global Burden of Disease Study. Lancet. 2017 Dec 2;390(10111):2437–60.

3. Budreviciute A, Damiati S, Sabir DK, Onder K, Schuller-Goetzburg P, Plakys G, et al. Management and Prevention Strategies for Non-communicable Diseases (NCDs) and Their Risk Factors. Vol. 8, Frontiers in Public Health. Frontiers Media S.A.; 2020. p. 788.

4. Al-Kuwari MG, Abdulmalik MA, Al-Mudahka HR, Bakri AH, Al-Baker WA, Abushaikha SS, et al. The impact of COVID-19 pandemic on the preventive services in Qatar. J Public Health Res. 2021;10(1):1–4.

5. Bramer CA, Kimmins LM, Swanson R, Kuo J, Vranesich P, Jacques-Carroll LA, et al. Decline in child vaccination coverage during the COVID-19 pandemic —… Morbidity and Mortality Weekly Report. 2020 Jul 1;20(7):1930–1.

6. Moynihan R, Sanders S, Michaleff ZA, Scott AM, Clark J, To EJ, et al. Impact of COVID-19 pandemic on utilisation of healthcare services: a systematic review. BMJ Open. 2021 Mar 1;11(3):e045343.

7. Peters DH, Garg A, Bloom G, Walker DG, Brieger WR, Hafizur Rahman M. Poverty and access to health care in developing countries. Vol. 1136, Annals of the New York Academy of Sciences. Ann N Y Acad Sci; 2008. p. 161–71.

8. Gottret P, Schieber G. Health Financing Revisited: A Practitioner’s Guide. Graham S, Ross-Larson B, Rosen C, Baxter J, editors. Health Financing Revisited - A Practicioner’s Guide. Washington DC: The World Bank; 2006. 1–324 p.

9. WHO. Global Health Workforce statistics database [Internet]. World Health Organzation. 2020 [cited 2022 Mar 14]. Available from: https://www.who.int/data/gho/data/themes/topics/health-workforce

10. Wold Health Organization. The Impact of the COVID-19 Pandemic on Non-Communicable Disease Resources: Result of a Rapid Assessment. World Health Organization. 2020. 1–32 p.

11. Sahoo K, Kanungo S, Mahapatra P, Pati S. Non-communicable diseases care during COVID-19 pandemic: A mixed-method study in Khurda district of Odisha, India. Indian Journal of Medical Research [Internet]. 2021 May 1 [cited 2022 Mar 14];153(5):649–57. Available from: https://journals.lww.com/ijmr/Fulltext/2021/05000/Non_communicable_diseases_care_during_COVID_19.23.aspx

12. Devi R, Goodyear-Smith F, Subramaniam K, McCormack J, Calder A, Parag V, et al. The Impact of COVID-19 on the Care of Patients With Noncommunicable Diseases in Low- and Middle-Income Countries: An Online Survey of Patient Perspectives. J Patient Exp. 2021 Jul 26;8.

13. Mistry SK, Ali Armm, Yadav UN, Ghimire S, Hossain MB, Shuvo S Das, et al. Older adults with non-communicable chronic conditions and their health care access amid COVID-19 pandemic in Bangladesh: Findings from a cross-sectional study. PLoS One. 2021 Jul 1;16(7):e0255534.

14. Woolf SH, Chapman DA, Sabo RT, Zimmerman EB. Excess Deaths from COVID-19 and Other Causes in the US, March 1, 2020, to January 2, 2021. Vol. 325, JAMA - Journal of the American Medical Association. American Medical Association; 2021. p. 1786–9.

15. Dessie ZG, Zewotir T. Mortality-related risk factors of COVID-19: a systematic review and meta-analysis of 42 studies and 423,117 patients. BMC Infect Dis [Internet]. 2021 Dec 1 [cited 2022 Aug 23];21(1):1–28. Available from: https://bmcinfectdis.biomedcentral.com/articles/10.1186/s12879-021-06536-3

16. Pécout C, Pain E, Chekroun M, Champeix C, Kulak C, Prieto R, et al. Impact of the COVID-19 Pandemic on Patients Affected by Non-Communicable Diseases in Europe and in the USA. International Journal of Environmental Research and Public Health 2021, Vol 18, Page 6697 [Internet]. 2021 Jun 22 [cited 2022 Aug 23];18(13):6697. Available from: https://www.mdpi.com/1660-4601/18/13/6697/htm

17. Sharma GD, Mahendru M. Lives or livelihood: Insights from locked-down India due to COVID19. Social Sciences & Humanities Open. 2020 Jan 1;2(1):100036.

18. Garg S, Basu S, Rustagi R, Borle A. Primary health care facility preparedness for outpatient service provision during the COVID-19 Pandemic in India: Cross-sectional study. JMIR Public Health Surveill. 2020 Apr 1;6(2).

19. Ummer O, Scott K, Mohan D, Chakraborty A, Lefevre AE. Connecting the dots: Kerala’s use of digital technology during the COVID-19 response. BMJ Glob Health [Internet]. 2021 Jul 26 [cited 2022 Aug 31];6(Suppl 5):5355. Available from: /pmc/articles/PMC8300548/

20. Singh V, Sarbadhikari S, Jacob A, John O. Challenges in delivering primary care via telemedicine during COVID-19 pandemic in India: A review synthesis using systems approach. J Family Med Prim Care [Internet]. 2022 [cited 2022 Aug 31];11(6):2581. Available from: https://journals.lww.com/jfmpc/Fulltext/2022/06000/Challenges_in_delivering_primary_care_via.52.aspx

21. Rise N. The Role of Community Health Workers in the COVID-19 Response in the Caribbean, an exploratory study. Eur J Public Health [Internet]. 2021 Oct 20 [cited 2022 Sep 1];31(Supplement_3). Available from: https://academic.oup.com/eurpub/article/31/Supplement_3/ckab164.349/6405984

22. Bhaumik S, Moola S, Tyagi J, Nambiar D, Kakoti M. Community health workers for pandemic response: a rapid evidence synthesis. BMJ Glob Health [Internet]. 2020 Jun 1 [cited 2022 Sep 1];5(6):e002769. Available from: https://gh.bmj.com/content/5/6/e002769

23. Suarez-Lledo V, Alvarez-Galvez J. Prevalence of health misinformation on social media: Systematic review. Vol. 23, Journal of Medical Internet Research. JMIR Publications Inc.; 2021.

24. Swire-Thompson B, Lazer D. Public health and online misinformation: Challenges and recommendations [Internet]. Vol. 41, Annual Review of Public Health. Annual Reviews; 2019 [cited 2022 Mar 11]. p. 433–51. Available from: https://www.annualreviews.org/doi/abs/10.1146/annurev-publhealth-040119-094127

25. Okan O, Bollweg TM, Berens EM, Hurrelmann K, Bauer U, Schaeffer D. Coronavirus-related health literacy: A cross-sectional study in adults during the COVID-19 infodemic in Germany. Int J Environ Res Public Health [Internet]. 2020 Jul 30 [cited 2022 Mar 11];17(15):1–20. Available from: https://www.mdpi.com/1660-4601/17/15/5503/htm

26. Zhu Y, Jiang Y. The Four-Stages Strategies on Social Media to Cope with “Infodemic” and Repair Public Trust: Covid-19 Disinformation and Effectiveness of Government Intervention in China. In: Proceedings - 2020 IEEE International Conference on Intelligence and Security Informatics, ISI 2020. Institute of Electrical and Electronics Engineers Inc.; 2020.

27. Allen H, Wright BJ, Harding K, Broffman L. The role of stigma in access to health care for the poor. Milbank Quarterly. 2014;92(2):289–318.

28. Stangl AL, Earnshaw VA, Logie CH, Brakel W Van, Simbayi LC, Barré I, et al. The Health Stigma and Discrimination Framework : a global, crosscutting framework to inform research, intervention development, and policy on health-related stigmas. BMC Med. 2019;17(31).

29. Asadi-Aliabadi M, Tehrani-Banihashemi A, Moradi-Lakeh M. Stigma in COVID-19: A barrier to seek medical care and family support. Med J Islam Repub Iran. 2020 Oct 30;34(1):1–3.

30. Approaches to Reducing Stigma - Ending Discrimination Against People with Mental and Substance Use Disorders - NCBI Bookshelf. In: Ending Discrimination Against People with Mental and Substance Use Disorders: The Evidence for Stigma Change. 2016. p. 20.

