## Supplemental file 1- Verbatims for "Was access and quality of healthcare affected during COVID-19 pandemic? A qualitative enquiry into healthcare access for non-communicable diseases in Central India"

Supplementary data:

### Verbatims:

#### Unavailability of instruments:

Age/ F/ U:

…if sugar could also be done at home, that would be great….they used to check our sugar before like they check our BP. But the machines were taken back by -Name of the Hospital-. So, they could not check our sugar(s) (levels)…

#### Apathy of healthcare provider:

50/ F/ U+ 55/ M/ U

R1: they won’t give the medicine.

R2: yes, they won’t.

I: Oh!

R2: we do an auto from here to go there, auto driver takes 100 rupees…

I: Right…

R2: while going, while coming back, 100 rupees, 200 rupees, and they don’t give the medicine.

I: And you don’t even get the medicine?

R2: don’t even give it…

-----------------------

R2: The date he (doctor) will write, say 13th, so (you have to) come on 13th only.

I-Okay, if you go one day before, then they refuse it (giving medicine)?

R2: yes, no-

R1: they won’t give the medicine.

R2: yes, they won’t.

Attitude of the physician:

R2: Now, who will say that.

I-You didn’t say this?

R1: we-

R2: No. they scold us before (saying) anything. Stay far-stay-far-stay far…

R1: to listen, they aren’t ready to even listen…

Stigmatization:

I: So, for the patients of blood pressure or sugar was their commute affected or any other thing happened?

R: Yes. Doctors are also seeing them from distance, this is what they say “we were seated so far during the consultation” “did not take BP measurements properly, not even sugar but they are giving us medicines. So, if they are not checking us, medicines we can take, but if it affects our health negatively, so that would be more loss than gain...”

Availability of doctors:

I-As you were saying that you went to ~name~ PHC, did you encounter any problems? Like medicines-?

R2: yes, regarding medicines, it is like, doctor is not available always, only one doctor is there, the one who gives medicines

R: Yes. There were times when medicine was unavailable. So, in between people even went to private to buy medicines. After that now they are getting medicines from here...

I: Alright.

R: Medicines are available, but they are saying that “without checking how are they giving us medicines”.

I: was not being able to meet a doctor or not getting BP checked a problem for you?

R: yeah. I was scared that I am not able to meet a doctor or get my BP checked. But I had faith in God.

There was no doctor here, so, everyone got very troubled here, and they went to private. And many people did not have any work, so they did not have money for taking private medication. So, because of this, population in this are was a bit troubled.

I: So, the PHC was closed, and they didn’t even have money for medicines?

R: It wasn’t closed for long, but it was closed…

R: I got it checked from the private clinic.

I: was it expensive?

R: yeah. I had to pay 60 rupees.

I - During the lockdown last year, did you face any difficulty in getting health services for your illness?

R - There have been difficulties in buying medicines

#### Incorrect feeding practices:

R: Because, people are saying that increase your capacity, so that corona would not be able to attack you, (we) people are eating a lot, what I mean to say is they stay at home, they are consuming oily food, they are making delicacies and eating, so there is an effect of that, like their BP is increasing, they are eating oily food, they are eating salty food, they are cooking like that, there are holidays due to lockdown, everywhere, everyone is at home. they are not walking, they are not jogging or physical activity, so blood pressure would increase in such condition...

I: was this a difficulty for you?

R: obviously….i had no job and then these expenses like doctor’s fees etc…

Lack of proper information

I: you could not visit this PHC because you did not know if it were open?

R: yeah. I got to hear from someone that medicines are available at Janta quarters.

I: did you go there?

R: no. I was not sure if it was actually open or not..

Threat perception:

R-Like, we are better at home, there is disease there too (healthcare facility), we are better off at home itself. *laughs*

R: Yes, there have been barriers, they are scared, that “if we go, there would be corona…” so in these there have been a lot of barriers. They are afraid to go to the hospitals, and there are certain patients who aren’t even seen.

R: Yes, people could not go on time. They could not do because of threat of corona "if we go there may be a corona affected patient is already sitting over there, so it (the virus) will attack us, engulf us" so due to this reason they could not go, and some people could not go to collect medicines...

#### Travel restrictions:

R: patient will have to reach the facility. While going to 1250 hospital, police asked us to go back. They were not allowing us to even walk on the streets.

R: yeah sometimes I used to skip (medicines) for 4-8 days if there was barricading, we could not come. But still it was okay.

I: okay. Did anybody come to your house to give medicines?

R: no…

I: did somebody come to check your BP at home?....ASHA or anyone?

R: nobody came for anything…

I: did anybody visit you to measure BP or give medicines?

R: no. nobody comes to even ask how is the BP now, if I am taking medicines or not…

I: okay. Did the Government machinery (including ASHA, AWW, etc) provide you with any help to obtain healthcare during this lockdown?

R: no nothing like that..I just stood in queues at -hospital name- and if I got the prescription paper made then I was able to meet the doctor.

I: you did not try to get investigations done in private….?

R: hospitals were not taking any patients, so I did not get any investigations done.

R: not at all. I could just not get any investigations done as investigations were not being done here. BP was also not checked.

R: no during those 3 months I did not get anything (investigation) done.

R: I had problem in terms of conveyance. But I bought the medicines the doctor had given me. I still take those medicines for BP.

I-So, in PHC they say that stay afar… (social distancing)

R2: Yes!

Desired action: Availability of healthcare services:

R: So, it should be like that for medicines and people the hospitals must open all the time. They were open, and are open today as well, we didn’t know about such a pandemic.

Desired action: Availability of healthcare services:

R: I think they should keep the PHC open so that the patient can visit and take medicines.

Desired action: availability of doctors:

FLW:

R: So, it should be like that for medicines and people the hospitals must open all the time. They were open, and are open today as well, we didn’t know about such a pandemic.

Desired action: availability of doctors:

R (m/u) : doctors should be available….atleast for patients like us.

#### Desired action: line listing:

R (m/u) : like you are calling to inquire, likewise if somebody can call and ask. Obviously everybody cannot be contacted, but atleast the regular patients should be contacted and information should be taken from them. So that they can be given the correct guidance like when to visit the hospital, etc. It is difficult to enter -hospital name- hospital anyway.

#### Desired action: line listing:

FLW (f/u):

R: So, for that, as we were also earlier.... the patients of BP and other ailments, their survey shall be conducted, and we know that these patient requires medicine, so we can, for example, in our area there are 40 patients, so for these 40 patients, their names and mobile number can be taken, and we can get the medicines issued and deliver them to their homes. So, this problem will not progress, we can do this so it doesn't progress further...

#### Desired action: availability of public transport/identifier:

R (f/u) : even during lockdown people like us should be allowed to go for a walk..like maybe for half an hour or so…not everybody has the facility for exercising at home. So maybe they can issue a card or something, like the ones people use to travel during lockdown, which will allow us to take a walk. That will let us maintain our exercise for BP and sugar.

#### Desired action: public transport:

R (f/r) : public transport should not be stopped so that patients can visit their doctors easily. Conveyance was a big problem during the lockdown.

#### Desired action: necessary consumables:

R: For example, sanitizer. Sanitizer, mask and other arrangements should reach public. What they say is we do not have them so how will we give, like that. So, these are the problems what else...

#### Facilitator: FLW

(f/u) BP was also not checked. So I got it checked from the madam at anaganwadi in our locality. She came to our house and checked my BP.

(f/u) R: I used to send my son…and she used to come and check my BP. She used to check everybody’s BP in our locality…

FLW: (f/r):

Me and my daughter, she also goes with me, to write… we used to go and check people’s BP at times…

#### Availability of healthcare services:

(f/r) R: I did not face any difficulty. I took medicines from the Army hospital. I get my BP checked once a month usually. During lockdown I got it checked by a nearby doctor.

R: PHC was open only. Our area had no problem. I was able to come. I used to come on Wednesdays or Thursdays for the medicines.

#### Facilitator: investigation:

R: no I got my investigations done at -Name of Hospital-.

R: yeah I used to get all my investigations done. Whatever the doctors used to prescribe, I would get it done. I would get it done at -hospital name- only.

(f/u) R: yeah i was able to come regularly. Our locality was locked but we were careful. I came every 15 days.

#### Family support:

(f/r) R: No, there is not difficulty. Sons do it.

(f/r) R: we can measure BP and sugar at home by buying the particular machine. But I don’t know how to use it. My children could do it for me. They did ask me to buy the machines.

#### Facilitator: finances:

R: no. I took medicines from the market.

R: yes I used to buy medicines from the nearby medical shop….

I: and did you have any problems in getting investigations for diabetes?

R: I used to measure my sugar at home….apart from this I did not need any other investigations….

#### Facilitator: home-delivery of medicines:

(FLW/u) R: Yes. I have done it, at time we get them medicines from 1100 (PHC/dispensary), at times there are elderly women who cannot go, so we...

(FLW/u) R: We have done it before as well, the elder ladies, who unable to go to health centers... We bring them tablets…

#### Facilitator: private clinics:

R: I got it checked every month in private only..

R: she did not check my sugar. I got it checked outside in private. I used to go at night and get it checked.

R: I went only because once this PHC was closed. It was not difficult as it was very cheap…14 rupees only…

Perception about healthcare professional’s perception:

R: Only because of corona, because they might also be scared that “why we should touch him?” that might also happen.

#### Incorrect health practices/information:

R: I did not face any difficulties as during lockdown I used to follow the diet of -name-. He is on youtube. I did not need much medicines. My sugar was also normal. I only required one tablet.

R: like If I had to take 2 tablets per day I took 1 only. It lasted me for 4-5 months. Then lockdown opened.
